# GMAC: A simple measure to quantify upper limb use from wrist-worn accelerometers

**DOI:** 10.1101/2023.11.26.23299036

**Authors:** Sivakumar Balasubramanian

## Abstract

Various measures have been proposed to quantify upper-limb use through wrist-worn inertial measurement units. The two most popular traditional measures of upper-limb use – thresholded activity counts (TAC) and the gross movement (GM) score suffer from high sensitivity and specificity, respectively. We had previously proposed a hybrid version of these two measures – the GMAC – that showed better overall detection performance. However, the previously proposed GMAC used both accelerometer and gyroscope data and used the same parameter values from the TAC and GM measures. In this paper, we aim to answer two important questions to improve the usefulness of the GMAC measure: (a) can the GMAC measure be implemented using only the accelerometer data? (b) what are the optimal parameter values for the GMAC measure? We propose a modified version of the GMAC that works with only accelerometer data, and optimize this measure’s parameters. This optimized GMAC showed better detection performance than the previously proposed GMAC and surprisingly had comparable performance to that of the best-performing machine learning-based measure (random forest inter-subject model). Although intra-subject machine learning-based measures perform better than the optimized GMAC, the latter is simpler, well suited for real-time upper-limb use detection, and is the best option when a trained machine learning-based intra-subject model or labeled data is unavailable. The optimized GMAC measure can be a useful measure for either offline detection or for real-time detection and feedback of upper limb use.

## 1 Introduction

There is a growing interest in using wearable sensors for tracking upper limb (UL) movement behavior outside the clinic to quantify participation [1–3]. This interest is fuelled by the need to go beyond conventional measures that rely on self-reported questionnaires and patient interviews [4], which lack objectivity and sensitivity. An ideal system for performing this assessment must consist of (a) an unobtrusive measurement system that seamlessly records movement-related information, and (b) an automated data analytics pipeline. Such a system can provide accurate and reliable quantitative answers to clinically relevant questions, such as, how much the two ULs are used, how symmetric is their use, how “good” are the movements, etc.

MEMS-based inertial measurement unit (IMU) is a compact, wearable sensor that measures linear acceleration and angular velocity of the rigid body to which it is attached. To track UL movements in daily life, the minimum number of sensors, ideally one sensor per limb, is preferred. The most popular choice for sensor location for this application is around the distal forearm just proximal to the wrist joint [4–8], since, (a) the forearm’s linear and angular kinematics are sensitive to both shoulder and elbow movements, (b) this location has the largest moment arm about the shoulder and elbow joints, thus, registering relatively large linear acceleration signals resulting from shoulder/elbow joint rotations, and (c) the ease of donning/doffing the sensor on this location,

The most fundamental construct of interest in UL functioning is *upper limb use* [4]. This is a binary construct indicating the presence or absence of a voluntary, meaningful UL movement or posture [9]. An accurate estimation of this complex construct requires access to information about the complete UL kinematics and kinetics, and the context in which the movement/posture is performed. In practice, a single wrist-worn IMU only provides the linear acceleration **a**_*S*_ and angular velocities of the ***ω***_*S*_ of the forearm in the local sensor reference frame, which is problematic for multiple reasons: (a) it cannot dissociate useful shoulder-elbow movements from unwanted movements, such as whole body movements, (b) it cannot detect finger movements, (c) it cannot ascertain if a movement/posture is voluntary, and (d) it is devoid of contextual information.

Nevertheless, several measures have been proposed in the literature to detect UL use from a single IMU [5, 7, 8, 10]. These measures can be broadly categorized into traditional [5, 10, 11] and machine learning(ML)-based measures [6,10]; we use the terms measure, model, and algorithm interchangeably in the rest of the manuscript. The traditional measures are simple, hand-crafted algorithms with pre-specified parameter values that use specific signal features to detect UL use. For instance, the thresholded activity counts (TAC) measure [5] uses the magnitude of the gravity-subtracted acceleration, while the gross movement (GM) score [4, 8] uses the orientation of the forearm and the amount of forearm movement. On the contrary, ML-based measures are algorithms trained on a set of labeled data to detect UL use from IMUs. Random forests, support vector machines, and multilayer perceptrons have been reported previously [6, 10], with the random forests [6, 10] offering the best performance to date. Additionally, intra-subject (i.e. subject-specific) ML models perform better than inter-subject (i.e. one model trained across different subjects) models [6, 10]. Although the ML-based measures perform better than the traditional methods, the latter has some advantages, such as:(a)they are simple and easy to interpret, and (b) they can be implemented efficiently in firmware for real-time detection and feedback of UL use (e.g. like the step count feedback from pedometers).

The TAC and the GM measures are the two most popular measures for quantifying UL use. Previous studies have shown that the TAC is a highly sensitive measure, while the GM is highly specific [10, 12]. We recently proposed a hybrid measure, called the *GMAC*, that combines the TAC and GM measures to balance out their respective high sensitivity and specificity [10]. The GMAC showed a better overall performance than TAC or GM, as quantified using the Youden index, but had a lower performance than the inter- and intra-subject ML measures. Our previous work had also shown that the best-performing ML measures used the mean and variance of the accelerometer signal to detect UL use; interestingly, the accelerometer’s mean and variance are related to the orientation of the forearm (used by the GM), and the variance is related to the amount of forearm movement (related to the GM and TAC). Thus, in principle, the GMAC and ML measures use similar information but different decision boundaries for deciding UL use. Given, that the GMAC is a simple and reasonable alternative to ML measures, a more detailed investigation of the GMAC algorithm and the optimization of its parameters to work effectively for both healthy and hemiparetic subjects are warranted. Thus, the aim of this study was to find answers two important questions about the GMAC algorithm:

1. Can the GMAC algorithm be implemented using only a wrist-worn accelerometer? This is an important question because: (a) some popular wearable sensors (e.g. from ActiGraph, USA) only contain an accelerometer, (b) gyroscopes do not add any values to UL use detection [10], and (c) gyroscopes are power-hungry sensors, and avoiding them can result in more efficient UL use trackers.
2. What are the optimal parameters for the GMAC algorithm that work well for both healthy and hemiparetic subjects? The parameters of the GMAC algorithm were previously chosen based on the TAC and GM, which might not be optimal.

The paper starts with a description of the GMAC algorithm proposed by Subash et al. [10], followed by the description of the newly proposed GMAC that works only with accelerometer data. The different parameters of this new algorithm and the estimation of their optimal values are then presented. The paper ends with a discussion of its results, its implications for clinical use, and the limitations of the current study.

## 2 Methods

This work uses data from our previous study [12] which is openly available as part of a Github repository. The data was collected from 10 healthy healthy subjects and 5 hemiparetic subjects, using a custom-built wearable IMU sensor that samples accelerometer and gyroscope data at 50Hz. The subjects performed various tabletop and non-tabletop tasks while wearing the IMU sensors on both forearms, proximal to the wrist joints. The tasks were video recorded simultaneously for labeling UL use employing the Functional Arm Activity Behavioural Observation System framework (FAABOS) [13]. More details about the data and the protocol can be found in [10, 12].

### 2.1 The previous GMAC measure

The accelerometer and gyroscope signals are given by **a**_*S*_ [*n*] and ***ω***_*S*_ [*n*], respectively, at the sampling time instant *n* ∈ ℤ; both signals are sampled at *f*_*s*_ = 50Hz. The previously proposed GMAC measure computes the UL use 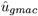 every second (i.e. every *f*_*s*_ samples) using the activity counts *α*_*vm*_ obtained from the vector magnitude algorithm [5], and the mean pitch angle 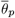 of the forearm,

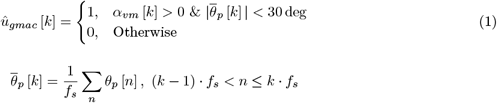

where, *k* ∈ ℤ represents the sampling time instants of 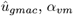 and 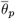, all of which are computed every one-second, *α*_*vm*_ [*k*] and 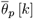 are the output of vector magnitude algorithm and the mean forearm pitch angle using the IMU data over the time window where (*k−*1) · *f*_*s*_ *< n≤ k*·*f*_*s*_, respectively. This previously reported algorithm uses both the accelerometer and the gyroscope signals to compute *α*_*vm*_ and 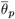 [10].

### 2.2 The new GMAC measure: using only the accelerometer data

If we only had the acceleration data **a**_*S*_ [*n*], we could still estimate information about the amount of forearm movement *α*_*gmac*_ [*n*] and forearm orientation *θ*_*gmac*_ [*n*]. A block diagram representation of the estimation procedure for *α*_*gmac*_ [*n*] and *θ*_*gmac*_ [*n*] from **a**_*S*_ [*n*] is shown in Figure 1, which also shows the various associated parameters with the different steps in these procedures (details in Table 1). The forearm orientation is computed as the *arccos* of the normalized component of the acceleration signal along the length of the forearm (which is taken as the *x* axis in Figure 1). The amount of forearm movement is computed by first highpass filtering the accelerometer data to remove the slow varying contribution from gravity, followed by computing the norm (Figure 1). Both of these signals are smoothed using moving average filters. The decision rule consists of two rules like Eq. 1 for detecting UL use,

**Table 1:**
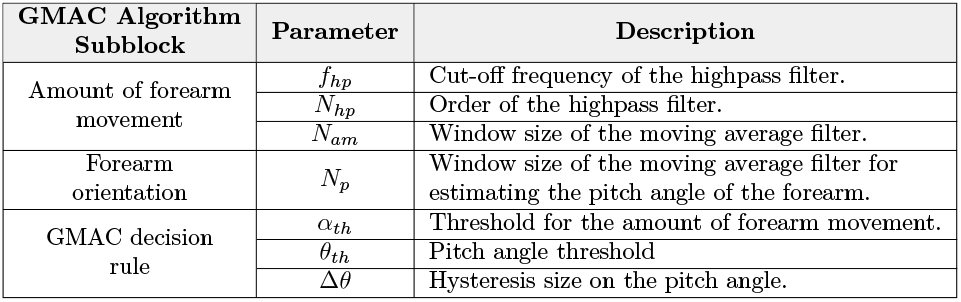
Description of the different parameters of the proposed GMAC algorithm depicted in Figure 1.

| GMAC Algorithm Subblock | Parameter | Description |
| --- | --- | --- |
| Amount of forearm movement | $f_{hp}$ | Cut-off frequency of the highpass filter. |
| | $N_{hp}$ | Order of the highpass filter. |
| | $N_{am}$ | Window size of the moving average filter. |
| Forearm orientation | $N_p$ | Window size of the moving average filter for estimating the pitch angle of the forearm. |
| GMAC decision rule | $\alpha_{th}$ | Threshold for the amount of forearm movement. |
| | $\theta_{th}$ | Pitch angle threshold |
| | $\Delta\theta$ | Hysteresis size on the pitch angle. |

**Figure 1:**
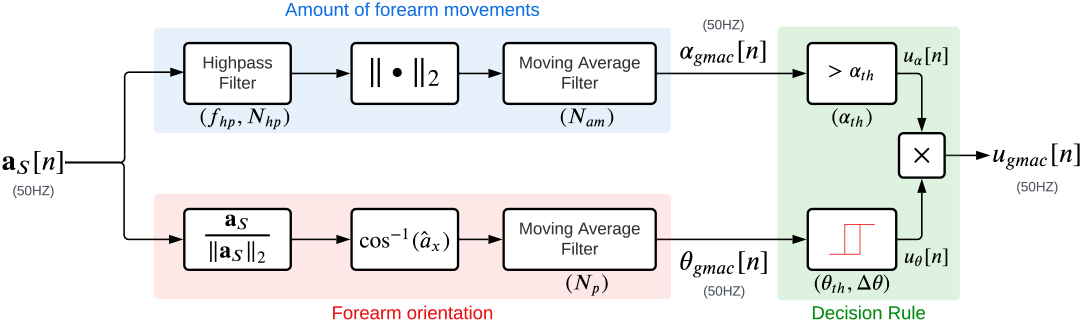
Schematic of the proposed GMAC algorithm to work with only accelerometer data. The proposed algorithm has three subblocks: (a) Forearm orientation (red background), (b) Amount of forearm movements (blue background), and (c) Decision rule (green background). The different parameters associated with the three blocks are shown in green colored text in the figure.

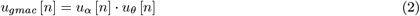

where, *u*_*α*_ [*n*], *u*_*θ*_ [*n*] ∈ { 0, 1} are obtained through thresholding rules applied on *α*_*gmac*_ [*n*] and *θ*_*gmac*_ [*n*], respectively, smilar to Eq. 1. The thresholding rule on *α*_*gmac*_ [*n*] is the following,

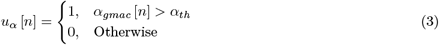

While, the second thresholding rule on *θ*_*gmac*_ [*n*] is a hysteresis rule, where the output at the time instant *n* depends on the current input *θ*_*gmac*_ [*n*] and the past value of the output *u*_*θ*_ [*n −* 1],

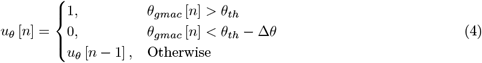

The choice of a simple thresholding rule for *α*_*gmac*_ [*n*] and a hysteresis rule for *θ*_*gmac*_ [*n*] was based on preliminary optimization work, indicating that a hysteresis rule on *α*_*gmac*_ [*n*] did not improve the detection performance. Eqs. 2, 3, and 4 constitute the new GMAC algorithm that uses only the accelerometer data **a**_*S*_ [*n*] to detect UL use. The optimization of the parameters associated with the different components of this algorithm is described in the next section.

### 2.3 Optimization of the new GMAC parameters

Instead of optimizing all seven parameters of the new GMAC algorithm to maximize detection performance, we chose a simpler approach where the parameters associated with the three subblocks (forearm orientation, amount of forearm movements, and decision rule) are optimized independently. The best parameters for the subblock for estimating the amount of forearm movement (*f*_*hp*_, *N*_*hp*_, *N*_*am*_) and the forearm orientation (*N*_*p*_) were first determined independently. These optimal parameters were then used to optimize the parameters of the decision rule (*α*_*th*_, *θ*_*th*_, Δ*θ*).

#### 2.3.1 Optimal parameters for estimating the amount of forearm movement

The optimal parameters were chosen as the set of parameters that maximized the Spearman correlation between *α*_*gmac*_ and the activity counts estimated from the vector magnitude algorithm *α*_*vm*_; the Spearman correlation was chosen because *α*_*gmac*_ and *α*_*vm*_ might be related nonlinearly. Given that *α*_*gmac*_ is computed at 50Hz, while the *α*_*vm*_ is computed at 1Hz, an equivalent variable was computed from *α*_*gmac*_ as the following,

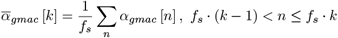

Both 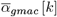 and *α*_*vm*_ [*k*] are both computed at 1Hz and the correlation between them was computed using the Spearman rank correlation coefficient. The correlation coefficients were computed for different values of the algorithm parameters by clubbing together data from both limbs of all healthy and hemiparetic subjects. A grid search for the parameter values was carried out over the ranges: *f*_*hp*_ *∈ {*0.01, 0.1, 1*}*Hz, *N*_*hp*_ *∈ {*2, 4*}*, and *N*_*am*_ *∈ {f*_*s*_, 5*f*_*s*_, 10*f*_*s*_*}*.

The effect of the different parameter values on the Spearman correlation coefficient was evaluated using a linear mixed-effects model. The three parameters were treated as fixed effects and the subjects were treated as a random effect; the statistical significance level was set at *p <* 0.05. The optimal parameter was chosen as the one with the highest median value for the correlation coefficient, across healthy and hemiparetic subjects for both limbs.

#### 2.3.2 Optimal parameters for estimating the forearm orientation

The optimal parameter value for *N*_*p*_ was chosen as the value that maximizes the Pearson correlation coefficient between the pitch angles estimates from the accelerometer *θ*_*gmac*_ [*n*] (Figure 1) and from the Madgwick algorithm using both the accelerometer and gyroscope *θ*_*p*_ [*n*]; these are expected to be linearly related. The correlation coefficient was computed separately for both the limbs from healthy and hemiparetic subjects for different values of the algorithm parameter *N*_*p*_ *∈ {*1, *f*_*s*_*/*2, *f*_*s*_, 2_*s*_, 4*f*_*s*_, 8*f*_*s*_*}*.

The effect of the different parameter values on the correlation coefficient was evaluated using a linear mixed-effects model. The parameter *N*_*p*_ was treated as a fixed effect and the subjects were treated as a random effect; the statistical significance level was set at *p <* 0.05. The optimal parameter was chosen as the one with the maximum correlation across healthy and hemiparetic subjects for both limbs.

#### 2.3.3 Optimal parameters for GMAC decision rule

The best parameters chosen from the previous two sections were then used to compute *α*_*gmac*_ [*n*] and *θ*_*gmac*_ [*n*], which were used to optimize the parameters (*α*_*th*_, *θ*_*th*_, Δ*θ*) of the decision rule. For each parameter combination, the Youden index [14] was computed for both limbs for all subjects (healthy and hemiparetic), as the following:

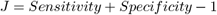

where the sensitivity and specificity are computed from the confusion matrix generated from the UL use detected using the new GMAC algorithm for the given parameter combination *u*_*gmac*_ (Figure 1) and the ground truth obtained from the FAABOS framework.

The optimal parameter combination for the decision rule was defined as the one that maximizes the overall detection accuracy, consistently (in terms of the Youden index) across both limbs for healthy and hemiparetic subjects. This was defined as the following optimization problem,

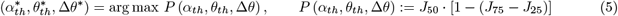

where 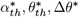 are the optimum parameter values, *J*_*q*_ is the *q*^*th*^ percentile of the Youden index computed for a given parameter combination for the two limbs for healthy and hemiparetic subjects. The median Youden index *J*_50_ in *P* is a measure of the detection accuracy, while the term [1*−* (*J*_75_*− J*_25_)] is a measure of the consistency of the detection accuracy of the given parameter combination, across the two limbs of healthy and hemiparetic subjects. A grid search for the parameter values was carried out over the ranges: *α*_*th*_ *∈ {*0, 0.1, 0.25, 0.5*}, θ*_*th*_ *∈ {−*90, *−*80, …, 90*}* deg, and Δ*θ ∈ {*0, 20, 40, 60, 80*}* deg.

An estimate of the expected performance of this process of optimizing the new GMAC measure parameters was computed by employing a leave-one-subject-out cross-validation approach. We have a total of 30 independent datasets (15 subjects and two limbs). The grid search optimization process was performed on 29 datasets to find the optimal parameters using the aforementioned process. This optimal parameter combination was then used to evaluate the detection performance on the one dataset that was excluded. This resulted in 30 Youden indices that provide an estimate of the expected performance of the new optimized GMAC measure on unseen data. This expected performance was compared against the Youden indices of the old GMAC, inter-subject random forest (RF Inter), and intra-subject random forest (RF Intra) measures from our previous work [10], through paired t-tests.

## 3 Results

All analyses in this work were carried out in Python using the Jupyter Notebook environment [15] and the linear mixed-effects modeling was performed using the ‘statsmodels’ package [16]. The plot of the effect of the parameter *N*_*p*_ on the estimation of the forearm orientation is shown in Figure 2(A). A linear mixed-effects model revealed a significant main effect due to *N*_*p*_ (*p ≪* 0.05). Based on these results, the optimal parameter for computing *θ*_*gmac*_ [*n*] was selected as 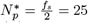, which showed the highest median Pearson correlation coefficients (Figure 2(A)). Figure 2(B)-(C) display the Spearman correlation between 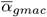 and *α*_*vm*_ as a function of the parameters *f*_*hp*_, *N*_*hp*_, and *N*_*am*_. A linear mixed-effects model revealed: (a) a significant main effect due to *f*_*hp*_ (*p ≪* 0.05) and (b) no significant main effect due to *N*_*hp*_; and (c) a significant main effect due to *N*_*am*_ (*p ≪* 0.05). Based on these results, the optimal parameters for computing *α*_*gmac*_ [*n*] were selected as: (a) 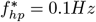 and 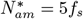 since these parameter values showed the highest median values for the correlation; 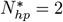 was chosen for a simpler filter structure.

**Figure 2:**
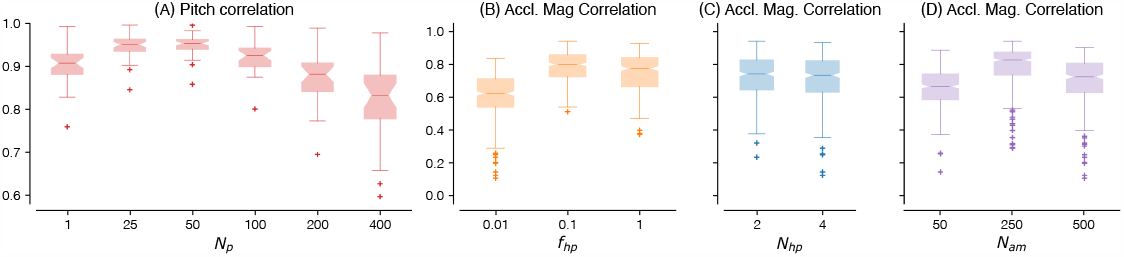
Boxplot depicting the performance of forearm orientation and amount of forearm movements estimation as a function of the different algorithm parameters. The median Spearman correlation coefficient for (A) forearm pitch estimation as a function of the parameter *N*_*p*_, and (B)-(D) acceleration magnitude estimated from Figure 1 and the vector magnitude algorithm [5] as a function of *N*_*c*_, *f*_*c*_, and *N*_*am*_, respectively, are shown in the plots.

The heatmaps of the performance measure *P* (*α*_*th*_, *θ*_*th*_, Δ*θ*) for both limbs of the healthy and hemiparetic subjects, as a function of *θ*_*th*_ and Δ*θ* for different values of *α*_*th*_ are shown in Figure 3. The darker regions of the heatmap correspond to higher performance; the maximum value of *P* and the corresponding values for the three parameters are depicted using a white square marker in the second heatmap from the left in Figure 3. The best parameters values identified through this procedure where 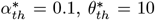, and Δ*θ*^*\**^ = 40. Only two other parameter combinations were in the top 5% of performance values, which are depicted using yellow circles in Figure 3. These two other parameter combinations were (*α*_*th*_ = 0., *θ*_*th*_ = 10°, Δ*θ* = 60°) and (*α*_*th*_ = 0.1, *θ*_*th*_ = 20°, Δ*θ* = 60°).

**Figure 3:**
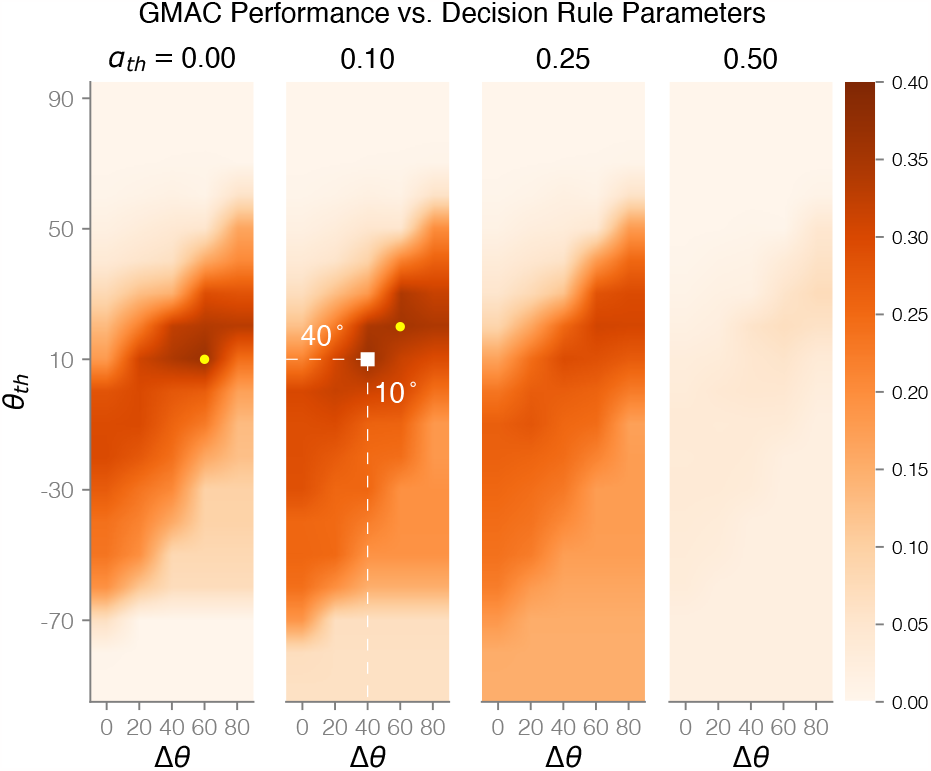
Heatmaps of the performance *P* (*α*_*th*_, *θ*_*th*_, Δ*θ*) as a function of the decision rule parameters. The individual heatmaps depict the performance as a function of *θ*_*th*_ and Δ*θ* for four different values of *α*_*th*_. The highest value of *P* (*α*_*th*_, *θ*_*th*_, Δ*θ*) is shown using a white square in the second heatmap from the left, which corresponds to the parameter values: *θ*_*th*_ = 10°, Δ*θ* = 40°, *α*_*th*_ = 0.1. The two yellow circles are two other parameter combinations whose performance values are within 5% of the maximum performance value: (*α*_*th*_ = 0., *θ*_*th*_ = 10°, Δ*θ* = 60°) and (*α*_*th*_ = 0.1, *θ*_*th*_ = 20°, Δ*θ* = 60°).

Figure 4 shows the comparison of the Youden index and the receiver operating characteristics plot of the optimized GMAC algorithm with that of the three different measures investigated by Subash et al. [10]: the original GMAC (Old-GMAC), the random forest inter-subject (RF-Inter) and intra-subject (RF-Intra) models. The left plot shows the Youden indices for both limbs of healthy and hemiparetic subjects; the Youden index for the optimized GMAC shown was computed using the leave-one-subject-out cross-validation. This plot depicts the bootstrap estimates of the mean and its 95% confidence interval for the Youden index for the different measures. The mean differences in the Youden index, sensitivity, and specificity between the optimized GMAC and the other three measures are shown in Table 2. Paired t-tests comparing the optimized GMAC with the measures from Subash et al. 2022 revealed that:

**Table 2:** Mean difference in the Youden index, sensitivity, and specificity between the optimized GMAC and the three measures investigated by Subash et al. 2022 [10]; ‘n.s.’ stands for ‘not significant.’

| Comparison with Subash et al. 2022 | Difference in Youden index | Difference in Sensitivity | Difference in Specificity |
| --- | --- | --- | --- |
| Old-GMAC | $0.097 \pm 0.130$ | $0.094 \pm 0.191$ | $0.003 \pm 0.217^{\text{n.s.}}$ |
| RF Inter-subject | $-0.043 \pm 0.143^{\text{n.s.}}$ | $-0.020 \pm 0.203^{\text{n.s.}}$ | $-0.024 \pm 0.213^{\text{n.s.}}$ |
| RF Intra-subject | $-0.286 \pm 0.224$ | $-0.064 \pm 0.238$ | $-0.222 \pm 0.259$ |

**Figure 4:**
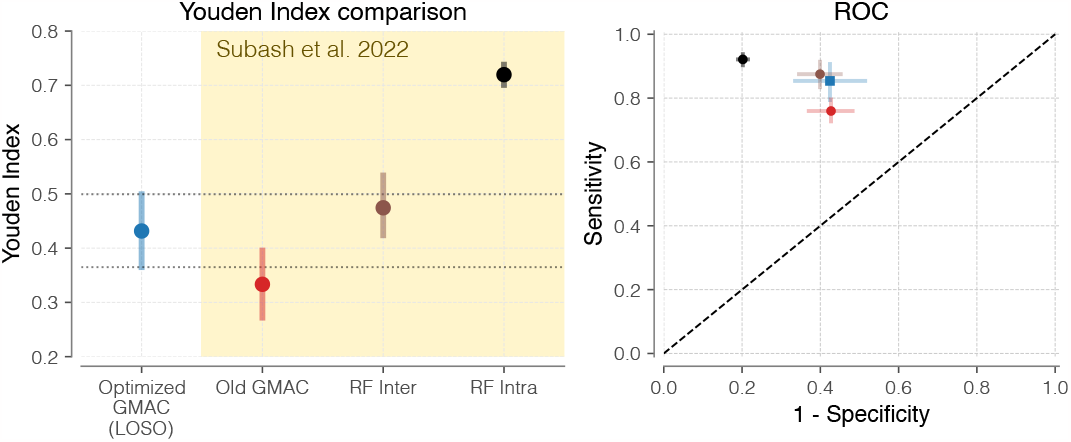
Comparison of four different measures of UL use: the optimized GMAC proposed in the current study, the old GMAC algorithm, random forest inter-subject (RF Inter), and random forest intra-subject (RF Intra) measures from Subash et al. [10]. The Youden index of the optimized GMAC algorithm obtained using the leave-one-subject-out cross-validation procedure is shown in the blue box plot, and that of the old GMAC, RF Inter, and RF Intra in orange, brown, and gray boxplots. The right plot shows the receiver operating characteristics plot for these four measures in their corresponding colors.

1. The optimized GMAC measure has significantly greater Youden index (*p <* 0.05) (Table 2) than the Old-GMAC measure proposed by Subash et al. 2022 [10]. This increased Youden index is due to increased sensitivity (Table 2). This is depicted in the receiver operating characteristic plot on the right of Figure 4.
2. The optimized GMAC measure is not significantly different from the RF-Inter measure from [10], which is an unexpected result.
3. The optimized GMAC measure is significantly worse than the RF-Intra measure from [10]. This result is expected since the RF-Intra model did not have to deal with inter-subject variability for detecting UL use [10].

## 4 Discussion

This work demonstrates that the GMAC measure can be computed from the accelerometer data and with the appropriate parameters this optimized GMAC measure performs better than the previously proposed GMAC measure; the optimized GMAC achieves significantly higher sensitivity without compromising on its specificity (Table 2). Surprisingly, this optimized GMAC measure performs as well as the random forest inter-subject model investigated by Subash et al. [10]; the two measures have similar sensitivities and specificities (Figure 4 and Table 2). The optimized GMAC’s performance, however, remains significantly lower than that of the random forest intra-subject model [10]. The results of this study have some important implications for the UL use detection problem and its use in clinical research and practice.

### 4.1 Why does the optimized GMAC perform better?

Given that the parameters of the GMAC measure were optimized in this study, it is reasonable that the optimized GMAC performs better than the previously proposed GMAC measure from [10]; Subash et al. used the same parameter values employed by the TAC and GM measure in their study [10]. It has been previously pointed out that the GM pitch angle thresholds of *±*30° might be conservative, as many functional movements require one to lift his/her forearm by more than +30° [12]. This range of *±*30° was originally proposed by Leuenberger et al. [8] was based only on visual observations of reaching movements and ADL. The formulation of the GMAC proposed in the current study addresses this issue by making all pitch angles greater than *θ*_*th*_ to be marked as functional if it satisfies the acceleration magnitude criterion. This could have helped improve the sensitivity compared to the previous GMAC measure. Another reason for the improved performance of the optimized GMAC could be the use of the hysteresis rule on the forearm pitch angle instead of the simple rule |*pitch*| *<* +30° (Eq. 1).

The optimized GMAC presented in this study is an inter-subject model – one that is optimized to work with either limb for healthy and hemiparetic subjects. However, it’s unclear why the optimized GMAC performs as well as the random forest inter-subject model from [10]. This was an unexpected result. A preliminary analysis involving the limb- and subject-wise optimization of the GMAC parameters (an “intra-subject” GMAC) revealed that the random forest intra-subject model was superior that the limb/subject-wise optmized GMAC (results not shown). One possible explanation for this is that the individual subject behavior is complex, whereas the average population-level behavior is simpler. The flexibility of the random forest intra-subject model allows it to learn this complex behavior at the subject level, while the relatively simple structure of the GMAC does not. This also explains the similar performance between the random forest inter-subject model and the optimized GMAC at the population level.

The optimal values for the decision rule parameter were chosen as 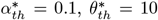, and Δ*θ*^*\**^ = 40. However, two other parameter combinations resulted in performance value that was within 5% of this best performance value: (*α*_*th*_ = 0., *θ*_*th*_ = 10°, Δ*θ* = 60°) and (*α*_*th*_ = 0.1, *θ*_*th*_ = 20°, Δ*θ* = 60°) (Figure 3). This suggests that the range of parameter values represented by these three combinations could provide similar detection performances.

### 4.2 Where will the GMAC be used?

The results of the current study indicate that the optimized GMAC is a superior alternative to existing traditional measures of UL use and the previously proposed by [10]. It is a simpler and easily implementable alternative to the random forest inter-subject model, which is currently the best-performing ML model for population-level data [6, 10].

However, intra-subject ML algorithms produce better UL use detection accuracy than traditional approaches [6, 10]. If available, optimal trained intra-subject models are currently the best option for offline UL use detection from previously recorded wearable sensor data. However, such ML-based algorithms may not be best suited for real-time UL use detection and feedback. The proposed GMAC measure (Figure 1) is an attractive alternative when, (a) trained ML intra-subject models are unavailable, (b) there is no annotated dataset to train new ML models, or (c) if real-time detection of UL use is required in an application. The GMAC measure can be efficiently implemented in the wearable sensor to detect UL use and intermittently transmit average UL use information to a mobile app for regular feedback. Given that the GMAC only involves simple linear filtering and thresholding rules (Figure 1), it is well suited for highly efficient firmware-level implementation in a wearable device. Future work must explore these possibilities of using GMAC to provide regular feedback to patients about UL use, which could encourage a hemiparetic subject to incorporate their affected limb in daily life.

### 4.3 Limitations of the current study

The current study has two main limitations. First, the data used in this study were collected from a small number of subjects (10 healthy and 5 hemiparetic subjects). The generalizability of the study results must be verified using a larger dataset, involving hemiparetic patients with a wide range of impairments, and by performing a larger subset of tasks. Second, the parameters of the GMAC algorithm were optimized in groups instead of optimizing them together. This type of optimization can result in suboptimal parameter values when there are interactions between parameters. However, this approach was chosen for its simplicity and future studies could explore better approaches than grip search for the parameter optimization [17].

## 5 Conclusion

The paper demonstrated how the GMAC can be derived from just the accelerometer data and showed that an optimized choice of the measure’s parameters leads to better performance than the old GMAC originally proposed by Subash et al. [10]. Surprisingly, the optimized GMAC had a similar performance to the random forest inter-subject measure [10], indicating that at the population level, the UL use behavior has a simple average structure. The proposed GMAC is a very attractive alternative when trained machine learning models are unavailable. The proposed GMAC algorithm can also be efficiently implemented in firmware for real-time detection and feedback of UL use, which is an important step towards encouraging UL use in hemiparetic patients. Future work involving a larger dataset, verifying the outcomes of the current study, exploring patient group-specific optimal GMAC measures, and efficient real-time implementation and evaluation are recommended.

## Data Availability

All data produced are available online at https://github.com/biorehab/upper-limb-use-assessment and https://github.com/siva82kb/gmac

https://github.com/siva82kb/gmac

## Acknowledgments

We would like to thank the members of the Biological Learning and Rehabilitation Group (CMC Vellore), Ms. Tanya Subash, and Dr. Sadhana Yadav for their critical comments and feedback about the manuscript.

## Data and Code Availability

The data used in the current study was made available as part of our previously published work in [10]. This data can be found at Upper-limb Assessment GitHub repository. The code used in the current study is available at GMAC GitHub repository.

## Conflict of Interest

The author declares no conflict of interest.

## Contribution

SB planned, designed, and executed the study, analyzed the data, and wrote the manuscript.

## Notes

### Competing Interest Statement

The authors have declared no competing interest.

### Funding Statement

This study did not receive any funding.

### Author Declarations

The study used ONLY openly available human data that were originally located at: https://github.com/biorehab/upper-limb-use-assessment

## References

[1] Aakash Kaku, Avinash Parnandi, Anita Venkatesan, Natasha Pandit, Heidi Schambra, and Carlos Fernandez-Granda. Towards data-driven stroke rehabilitation via wearable sensors and deep learning. In Proceedings of Machine Learning Research, volume 126, pages 143–171, 2020.

[2] Issam Boukhennoufa, Xiaojun Zhai, Victor Utti, Jo Jackson, and Klaus D. McDonald-Maier. Wearable sensors and machine learning in post-stroke rehabilitation assessment: A systematic review. Biomedical Signal Processing and Control, 71:103197, jan 2022.

[3] Dongni Johansson, Kristina Malmgren, and Margit Alt Murphy. Wearable sensors for clinical applications in epilepsy, Parkinson’s disease, and stroke: a mixed-methods systematic review. Journal of Neurology, 265(8):1740–1752, aug 2018.

[4] Ann David, Tanya Subash, S. K. M. Varadhan, Alejandro Melendez-Calderon, and Sivakumar Balasubramanian. A Framework for Sensor-Based Assessment of Upper-Limb Functioning in Hemiparesis. Frontiers in Human Neuroscience, 15, jul 2021.

[5] Ryan R. Bailey, Joseph W. Klaesner, and Catherine E. Lang. An accelerometry-based methodology for assessment of real-world bilateral upper extremity activity. PLoS ONE, 9(7), 2014.

[6] Peter S. Lum, Liqi Shu, Elaine M. Bochniewicz, Tan Tran, Lin Ching Chang, Jessica Barth, and Alexander W. Dromerick. Improving Accelerometry-Based Measurement of Functional Use of the Upper Extremity After Stroke: Machine Learning Versus Counts Threshold Method. Neurorehabilitation and Neural Repair, 34(12):1078–1087, 2020.

[7] Diogo S. De Lucena, Oliver Stoller, Justin B. Rowe, Vicky Chan, and David J. Reinkensmeyer. Wearable sensing for rehabilitation after stroke: Bimanual jerk asymmetry encodes unique information about the variability of upper extremity recovery. IEEE International Conference on Rehabilitation Robotics, pages 1603–1608, 2017.

[8] Kaspar Leuenberger, Roman Gonzenbach, Susanne Wachter, Andreas Luft, and Roger Gassert. A method to qualitatively assess arm use in stroke survivors in the home environment. Medical and Biological Engineering and Computing, 2017.

[9] Neville Hogan and Dagmar Sternad. On rhythmic and discrete movements: Reflections, definitions and implications for motor control. Experimental Brain Research, 181(1):13–30, jun 2007.

[10] Tanya Subash, Ann David, StephenSukumaran ReetaJanetSurekha, Sankaralingam Gayathri, Selvaraj Samuelkamaleshkumar, Henry Prakash Magimairaj, Nebojsa Malesevic, Christian Antfolk, Varadhan SKM, Alejandro Melendez-Calderon, and Sivakumar Balasubramanian. Comparing algorithms for assessing upper limb use with inertial measurement units. Frontiers in Physiology, 13, ec 2022.

[11] G. Uswatte, W. H. R. Miltner, B. Foo, M. Varma, S. Moran, and E. Taub. Objective Measurement of Functional Upper-Extremity Movement Using Accelerometer Recordings Transformed With a Threshold Filter. Stroke, 31(3):662–667, 2000.

[12] Ann David, StephenSukumaran ReethaJanetSureka, Sankaralingam Gayathri, Salai Jeyseelan Annamalai, Selvaraj Samuelkamleshkumar, Anju Kuruvilla, Henry Prakash Magimairaj, SKM Varadhan, and Sivakumar Balasubramanian. Quantification of the relative arm use in patients with hemiparesis using inertial measurement units. Journal of Rehabilitation and Assistive Technologies Engineering, 8:205566832110196, jan 2021.

[13] Gitendra Uswatte and Laura Hobbs Qadri. A Behavioral Observation System for Quantifying Arm Activity in Daily Life After Stroke. Rehabilitation Psychology, 2009.

[14] W. J. Youden. Index for rating diagnostic tests. Cancer, 3(1):32–35, 1950.

[15] Thomas Kluyver, Benjamin Ragan-Kelley, Fernando Pérez, Brian Granger, Matthias Bussonnier, Jonathan Frederic, Kyle Kelley, Jessica Hamrick, Jason Grout, Sylvain Corlay, Paul Ivanov, Damián Avila, Safia Abdalla, and Carol Willing. Jupyter Notebooks—a publishing format for reproducible computational workflows. In Positioning and Power in Academic Publishing: Players, Agents and Agendas - Proceedings of the 20th International Conference on Electronic Publishing, ELPUB 2016, pages 87–90, 2016.

[16] Skipper Seabold and Josef Perktold. Statsmodels: Econometric and Statistical Modeling with Python. In Proceedings of the 9th Python in Science Conference, pages 92–96, 2010.

[17] James Bergstra and Yoshua Bengio. Random search for hyper-parameter optimization. Journal of Machine Learning Research, 13:281–305, 2012.

